# Clonal hematopoiesis of indeterminate potential is associated with worse kidney function and anemia in a cohort of patients with advanced chronic kidney disease

**DOI:** 10.1101/2021.06.30.21259343

**Authors:** Caitlyn Vlasschaert, Amy J. M. McNaughton, Wilma Hopman, Bryan Kestenbaum, Cassianne Robinson-Cohen, Jocelyn Garland, Sarah M. Moran, Rachel Holden, Matthew B. Lanktree, Michael J. Rauh

**Author notes:** contributed equally (Matthew B. Lanktree, Michael J. Rauh).

## Abstract

**Background:** Clonal hematopoiesis of indeterminate potential (CHIP) is an inflammatory premalignant disorder resulting from acquired genetic mutations in hematopoietic stem cells. CHIP is common in aging populations and associated with cardiovascular morbidity and overall mortality, but its role in chronic kidney disease (CKD) has not been investigated.

**Methods:** We performed targeted sequencing to detect CHIP mutations in a cohort of 87 adults with eGFR < 60 ml/min/1.73m^2^. Kidney function, hematologic, and mineral bone disease parameters were assessed cross-sectionally at baseline, and a total of 2,091 creatinine measurements and 3,382 hemoglobin measurements were retrospectively collected over the following 12-year period.

**Results:** At baseline, 20 of 87 (23%) cohort participants had CHIP detected. Those with CHIP had lower baseline eGFR (22.3 ± 11.2 vs. 28.2 ± 11.5 ml/min/1.73 m^2^, *P* = 0.04) in age- and sex-adjusted regression models. Individuals with CHIP had a 2.5–fold increased risk of incident 50% decline in eGFR or ESKD in a Cox proportional hazard model adjusted for age and sex (95% confidence interval, 1.3–4.7). The annualized rate of eGFR decline adjusted for age and sex was -2.3 ±1.1 ml/min/1.73m^2^ per year in those with CHIP *versus* -1.6 ±0.5 ml/min/1.73m^2^ per year in those without CHIP. Further, those with CHIP had lower hemoglobin at baseline (11.6 ± 0.3 vs. 12.8 ± 0.2 g/dL, *P* = 0.0003) and throughout the follow-up period despite a greater use of erythropoiesis-stimulating agents.

**Conclusion:** In those with pre-existing CKD, CHIP was associated with lower eGFR at baseline, faster progression of CKD, and anemia.

## Introduction

Clonal hematopoiesis of indeterminate potential (CHIP) is a newly recognized, age-associated hematologic disorder that imposes a significant risk of multi-system morbidity and mortality.^1^ When hematopoietic stem cells (HSCs) acquire mutations in key cell regulatory genes, biased production of a clonal myeloid cell population can result with altered immune functions.^2–5^ The three most commonly mutated genes driving CHIP – *TET2, DNMT3A* and *ASXL1* – are epigenetic regulators. A diagnosis of CHIP is made when the proportion of genetically identical progeny account for more than 4% of nucleated cells in the blood compartment. Based on this definition, the prevalence of CHIP is 10-20% in persons aged 65 and older^6,7^ and subthreshold clones are nearly ubiquitous by the fifth decade of life.^8^ Akin to other premalignant states, CHIP is associated with transformation to hematologic cancers; however such cancers are uncommon in affected persons with an estimated annual incidence of 0.5-1%.^9,10^ Strikingly, CHIP also portends a heightened risk of cardiovascular diseases and all-cause mortality.^11,12^ In cardiovascular disease, the risk conferred by CHIP is greater than many traditional cardiovascular risk factors including hypertension, smoking, and dyslipidemia.^13^

Myeloid cells carrying CHIP mutations produce high levels of inflammatory cytokines^6^, and both dysregulated interleukin-6 (IL-6) and inflammasome signaling have been causally implicated in CHIP pathogenesis.^2–5,14^ Moreover, experimental recapitulation of CHIP by transplanting only a small fraction of affected HSCs leads to kidney tubulointerstitial fibrosis and replacement of resident macrophages with clonal progeny.^4,15^ Since macrophages and neutrophils play pivotal roles in innate immunity, response to injury, management of the kidney interstitial environment, and inflammatory signalling, we hypothesized that persons harboring a CHIP mutation would have more rapid progression of CKD. There have been no published studies on CHIP in CKD. In this report, we analyzed associations of CHIP with kidney functional decline and its complications in a cohort of 87 individuals with baseline CKD followed from 2009 to 2020.

## Methods

### Study population

In 2009, 87 individuals from a single centre were enrolled in a longitudinal cohort study.^16–21^ Inclusion criteria included CKD stages G3–5 and age over 18 years with no prior history of hematologic malignancy. Participants had a median follow-up of 10.8 years [interquartile range: 5.3-11.4]. The study was approved by Queen’s University Health Sciences and Affiliated Teaching Hospitals Research Ethics Board (file #6023740).

### Measurement of the exposure

Peripheral venous blood was collected in PAXgene tubes at enrollment, DNA was extracted, and stored at –80°C. Genomic DNA (gDNA) was further purified using Axygen AxyPrep FragmetSelect I kit, then quantified using TaqMan RNaseP qPCR assay. DNA libraries were constructed using Ion AmpliSeq Kit for Chef DL8 with a validated custom AmpliSeq targeted CHIP 48-gene panel (genomic coverage as previously described^7^) on the Ion Chef instrument, followed by sequencing using an Ion 540 chip on the Ion S5XL sequencer (Thermo Fisher Scientific). The mean depth of coverage was >1,500× for all four sequencing runs, and the sample with lowest coverage had 1,409× coverage. Sequences were aligned to hg19 and variants were called using Ion Torrent Suite Software. Putative somatic mutations were identified using Ion Reporter software, excluding common SNPs in the University of California Santa Cruz (UCSC) database, and rare SNPs in the Genome Aggregation Database (gnomAD), filtering for exonic location, non-synonymous substitution, Phred quality score >20, coverage >50, VAF between 0.02 and 0.44, and visualizing in Integrated Genomics Viewer (IGV). The corresponding variant allele fraction (VAF) was recorded for each filtered variant. Participants were classified as having CHIP (CHIP+) if they had at least one CHIP driver mutation at a VAF ≥ 0.02, inferred to mean that the allele is present in a heterozygous state in ≥4% of diploid cells. Those that did not meet these criteria were classified as not having CHIP (CHIP–).

### Measurement of the outcome

The primary outcome was a sustained 50% decline in estimated GFR from the baseline value or end stage kidney disease, defined by an eGFR <15 ml/min/1.73m^2^, initiation of maintenance dialysis, or kidney transplantation. A sustained 50% decline in eGFR and a sustained eGFR <15 ml/min/1.73m^2^ required two consecutive qualifying measurements ≥28 days apart. The secondary outcome was the longitudinal slope of decline in eGFR.

We abstracted all serum creatinine (SCr) measurements from routine scheduled outpatient bloodwork from 2009 until December 31, 2020, excluding data from hospitalizations and “for cause” assessments. A total of 2,091 SCr measurements were collected over 527 years of follow-up for an average of four measurements per year per participant. SCr concentrations were measured using an isotope dilution mass-spectrometry traceable assay (Roche Creatinine Plus Modular assay). We calculated the estimated glomerular filtration rate (eGFR) using the CKD-Epidemiology Collaboration (EPI) 2009 equation.^22^ We ascertained dialysis, transplantation, and mortality status by electronic chart review.

### Measurement of other study data

We obtained clinical data, including medical histories and social habits, by participant self-report. We determined medication histories by electronic chart review, including prescriptions of erythropoiesis-stimulating agents (ESAs) and calcitriol. Synchronous with the time of DNA collection, we abstracted laboratory data including urea, bicarbonate, phosphate, calcium corrected for albumin, intact parathyroid hormone (PTH), complete blood count (CBC), C-reactive protein (CRP), and ferritin, as well as urine albumin–creatinine ratio (UACR).

Parameters of hematopoietic, inflammatory, and bone mineral status were also evaluated. Longitudinal hemoglobin data was averaged for every year of follow-up on a per-participant basis, and the average hemoglobin by CHIP status was assessed at baseline and at each year of follow-up, with censoring upon receipt of a kidney transplant when applicable. Rates of anemia, defined as hemoglobin < 12 g/dL for females and hemoglobin < 13 g/dL for males^23^, were also compared between those with and without CHIP.

### Statistical analysis

We tabulated baseline characteristics according to the presence *versus* absence of CHIP. For analyses of kidney disease progression, participants were followed from the ascertainment of CHIP status until they incurred this outcome or their data were censored due to death, loss to follow-up, or the end of the study period, whichever came first. We constructed Cox proportional-hazards regression models to estimate the association of CHIP status at baseline with the time to kidney disease progression with adjustment for age, sex, and baseline eGFR. To quantify the association of CHIP status with the slope of eGFR decline we used mixed model regression allowing for random slopes and intercepts of eGFR decline in each participant, while testing for a fixed effect of CHIP on eGFR across participants. For longitudinal analyses, we limited the observation period to the first three years after CHIP genotyping, where 41 (70%) participants remained uncensored to minimize non-random censoring due to death or initiation of dialysis.

Baseline laboratory data (kidney function, hematologic parameters, bone mineral markers, and inflammatory markers) were analyzed using linear regression by CHIP status and by quantitative VAF.

All analyses were performed using R statistical software version 4.0.5, with the significance level (α) set to 0.05.

## Results

### Prevalence of CHIP in the advanced CKD cohort at baseline

The mean age of the study population was 64 ± 14 years; 43% were women; and the mean baseline eGFR was 27 ± 11 ml/min/1.73m^2^.

Twenty of 87 individuals (23%) within the study population were found to carry CHIP variants at a VAF ≥2% (CHIP+, Figure 1). The most commonly affected genes were *TET2* (9/20), *DNMT3A* (8/20) and *ASXL1* (3/20). Five participants had multiple CHIP variants, and the average VAF for the largest detected clones per individual was 12.3%. Participants with CHIP were older and had a lower eGFR at baseline (Table 1).

**Figure 1 –.**
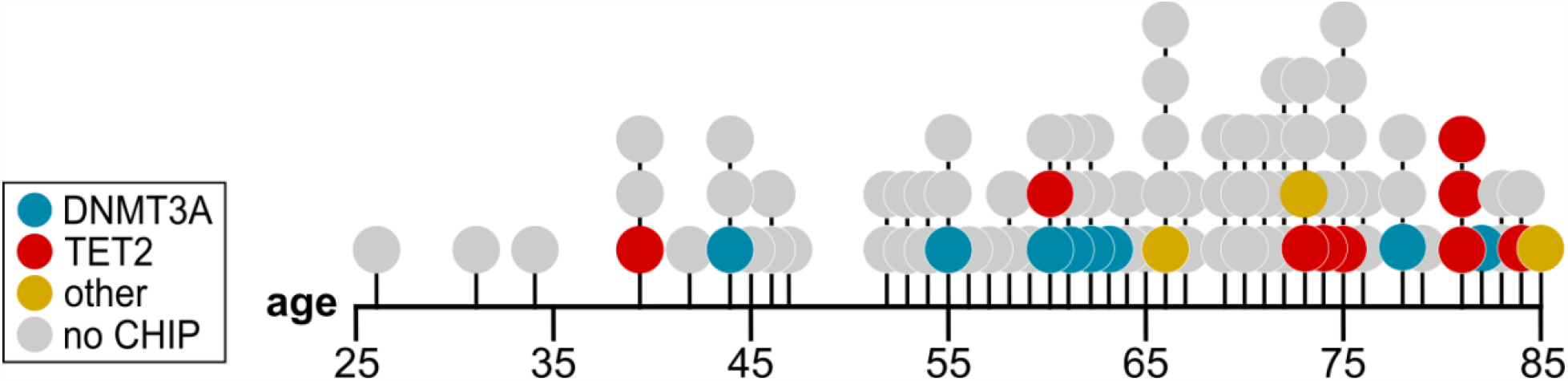
Age distribution of CHIP carriers in this cohort. The prevalence of CHIP increases with age and the genes affected are color-coded.

**Table 1.** Baseline cohort characteristics

| Characteristic | All (N = 87) | CHIP (N = 20) | No CHIP (N = 67) |
| --- | --- | --- | --- |
| Age | 63.9 ± 14.0 | 68.6 ± 14.3 | 62.5 ± 13.9 |
| Female | 43% | 45% | 43% |
| Baseline eGFR (CKD-EPI) | 26.9 ± 11.2 | 22.3 ± 11.2 | 28.2 ± 11.5 |
| Urine ACR | 75.0 ± 102.2 | 67.7 ± 64.5 | 77.4 ± 110.9 |
| Microalbuminuria (uACR ≥ 30) | 36 (41.4) | 10 (50.0) | 26 (38.8) |
| Macroalbuminuria (uACR ≥ 300) | 5 (5.7) | 0 (0) | 5 (7.5) |
| BMI (kg/m<sup>2</sup>) | 31.1 ± 7.0 | 31.4 ± 7.7 | 31.1 ± 6.8 |
| Hypertension<br>N (%) | 71 (93.1) | 19 (95.0) | 62 (92.5) |
| Diabetes<br>N (%) | 33 (37.9) | 6 (30.0) | 27 (40.3) |
| Smoking<br>N (%) | 48 (55.2) | 10 (50.0) | 38 (56.7) |
| Dyslipidemia<br>N (%) | 75 (86.2) | 16 (80.0) | 59 (88.1) |

### Association of CHIP with kidney disease progression

The primary outcome of a sustained 50% drop in eGFR or ESKD occurred in 14 persons with CHIP (incidence rate 22.8/100 person-years) and 29 persons without CHIP (incidence rate 10.3/100 person-years). After adjustment for age and sex, the presence of CHIP was associated with an estimated 2.5 times greater risk of kidney disease progression (95% CI 1.3 to 4.7 times greater risk; Figure 2C). This risk estimate was attenuated by additional adjustment for baseline eGFR (HR 1.8; 95% CI 0.9–3.4). The annualized rate of eGFR decline adjusted for age and sex was -2.3 ±1.1 ml/min/1.73m^2^ per year in those with CHIP *versus* -1.6 ±0.5 ml/min/1.73m^2^ per year in those without CHIP (Figure 2B).

**Figure 2 –.**
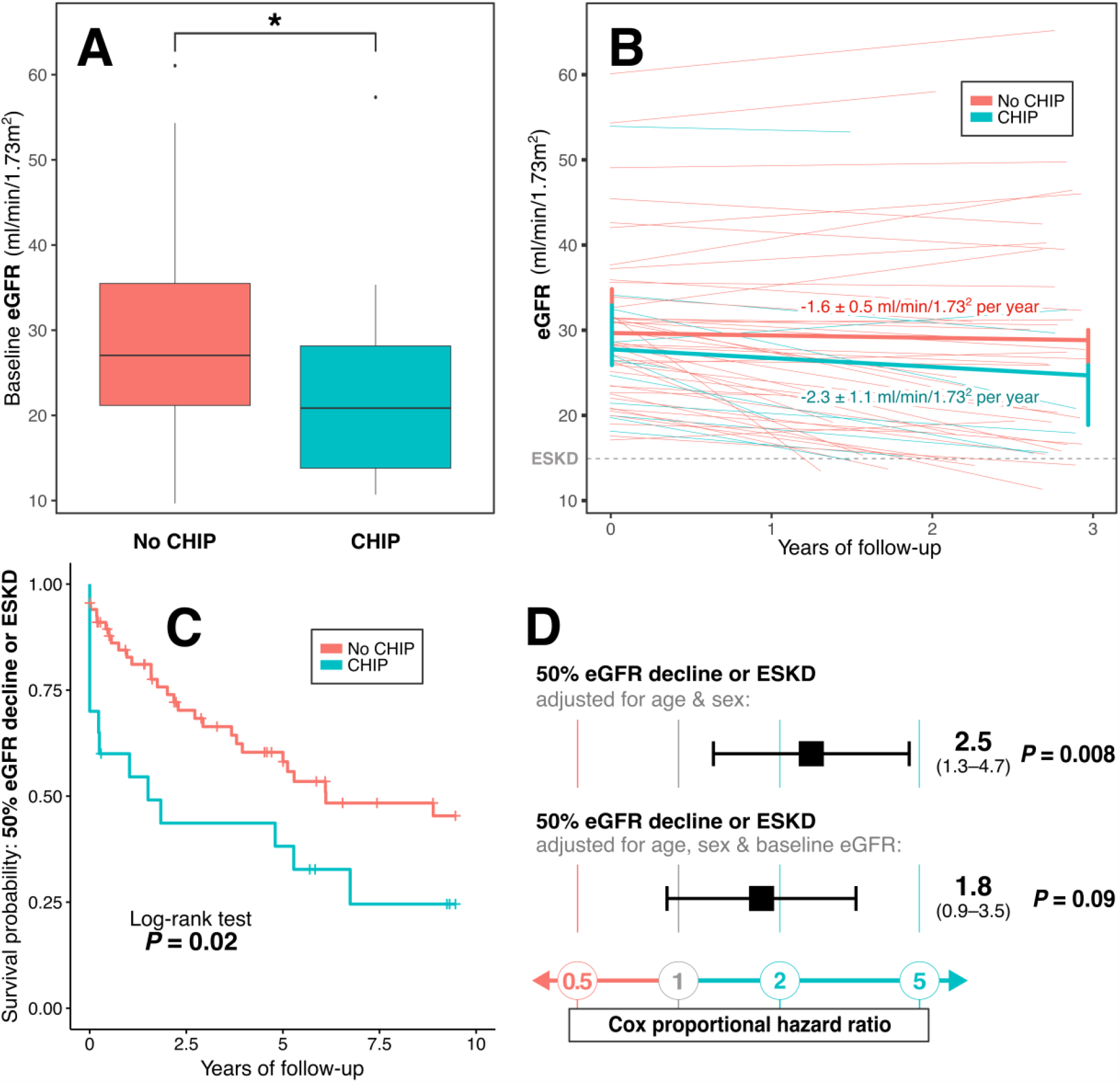
CHIP is associated with worse baseline kidney function and progression of CKD. A) The average baseline eGFR is significantly lower in those with CHIP compared to those without CHIP (N = 87). B) Mixed model regression of eGFR trends by CHIP status for the first three years of follow-up (see *Methods* for design rationale). Mixed model assumes CHIP, age and sex as fixed effects and allows random effects at baseline and along the y-axis. Only individuals who are pre-ESKD (eGFR > 15) and who have adequate follow-up (followed for >1 year and with >3 eGFR measurements) are included in the analysis (N = 59). Regression lines by dichotomous CHIP status and per individual are shown. C) Survival analysis for the composite outcome of 50% eGFR decline or ESKD by CHIP status (N = 87). D) Cox proportional hazard ratios for composite outcome of 50% eGFR decline or ESKD by CHIP status.

### CHIP and complications of CKD

We evaluated the association between CHIP and common complications of CKD including anemia, hyperparathyroidism, hyperphosphatemia, and metabolic acidosis. Participants with CHIP were more likely to be anemic at baseline (80% vs. 39% of individuals affected; **χ**^2^ = 10.5, *P* = 0.001). Hemoglobin concentration was significantly lower in those with CHIP at baseline (11.6 ± 0.3 vs. 12.8 ± 0.2 g/dL, *P* = 0.0003), and this significant difference between groups persisted over the entire follow-up period (Figure 3A). Even after adjusting for age, sex, and baseline eGFR, baseline hemoglobin was 1.2 ± 0.4 g/dL lower in those with CHIP (*P* = 0.002, Table 2). Ferritin levels were also significantly elevated in those with CHIP (250 ± 48 vs. 135 ± 14 µg/L), including after adjustment for age, sex, and baseline eGFR (Table 2, β = 101.5 ± 37.2 µg/L, *P* = 0.008), while transferrin saturation was within the normal range and not significantly different between groups (Figure 3C and Table 2). This suggests that iron deficiency is not a significant contributor to the observed anemia; in fact, the higher ferritin in those with CHIP may reflect greater inflammation. Ferritin levels were significantly negatively associated with hemoglobin levels (R = -0.405, *P* < 0.0001) while log-transformed C-reactive protein (log-CRP) levels were not significantly correlated with hemoglobin (R = -0.192, *P* = 0.08). The association between CHIP and lower hemoglobin remained when ferritin and log-CRP were introduced as independent covariates in the model (β = -0.8 ± 0.4 g/dL, *P* = 0.036). Altogether, CHIP status was significantly correlated with lower hemoglobin, but this was not fully explained by higher levels of inflammatory markers. It was also not explained by differences in the use of ESAs; in fact, those with CHIP were more frequently prescribed ESAs at baseline (40% vs 13% cross-sectional prevalence, *P* = 0.011) and during the follow-up period (70% vs. 42% cumulative incidence, *P* = 0.026; Figure 2B). The average mean corpuscular volume (MCV) was significantly associated with CHIP VAF in our cohort (Supplemental Table 2), which has been previously reported and attributed to mild defects in erythropoiesis in CHIP.^10^

**Figure 3 –.**
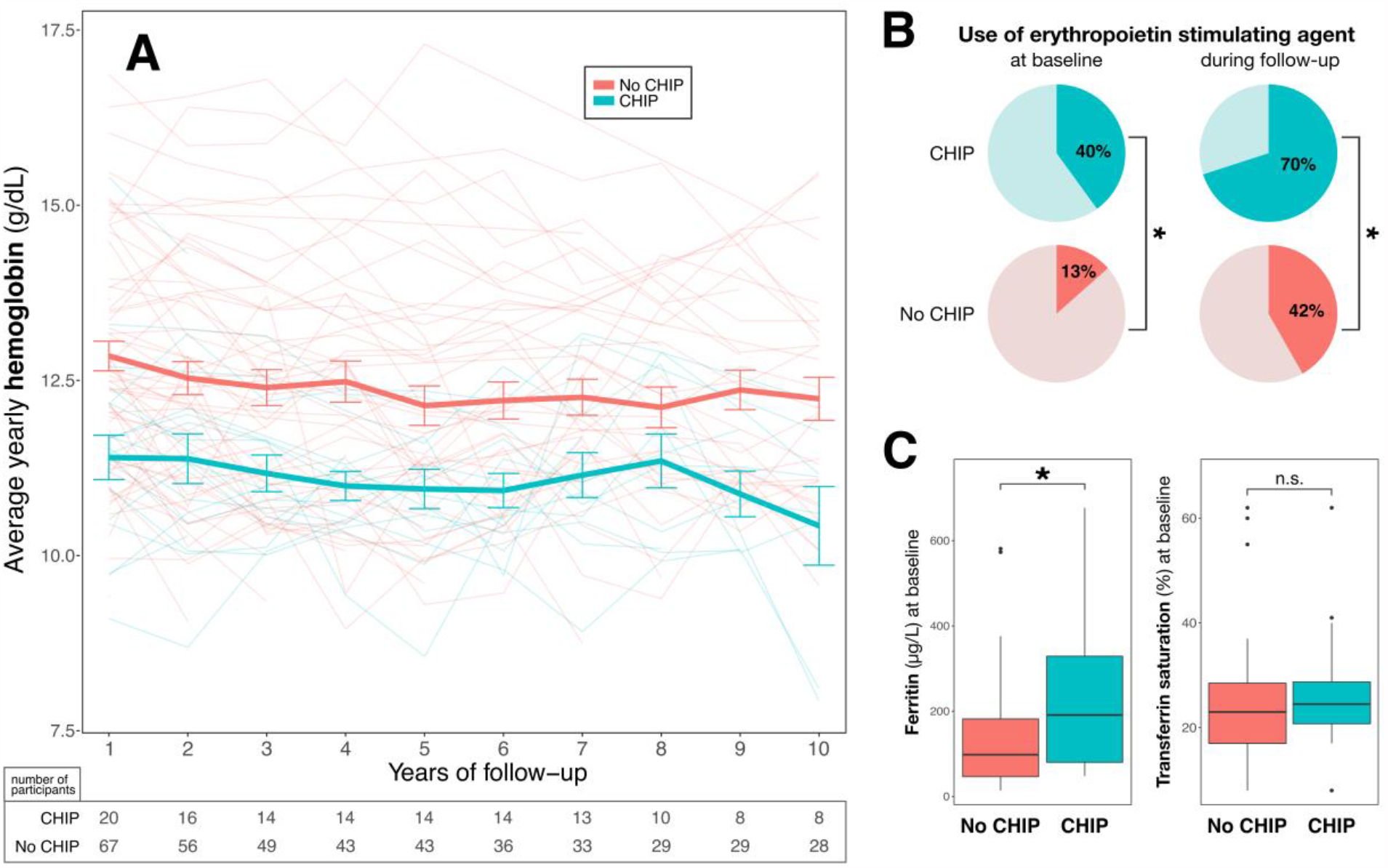
CHIP is associated with lower hemoglobin, higher ferritin and greater EPO use. A) Hemoglobin values– averaged per-person over yearly periods and shown with standard error (SE)– are significantly lower in those with CHIP throughout the follow-up period (N = 87). Hemoglobin values are censored upon receipt of a kidney transplant. B) The rate of erythropoiesis-stimulating agent (ESA) use was higher among those with CHIP, at cohort baseline and cumulatively during the follow-up period (N = 87). Asterisks (*) indicate statistically significant differences (p < 0.05) in the relative frequencies of ESA use between groups by chi-squared testing. C) Ferritin levels are significantly higher among those with CHIP on independent sample t-testing (p < 0.05), whereas transferrin saturation levels are not different (N = 87 for both).

**Table 2 –.** CHIP and CKD complications. The results of univariable (CHIP) and multivariable (CHIP, age, sex, baseline eGFR) linear regressions are presented for each outcome of interest. The β coefficient indicates the quantitative change in the outcome for those with CHIP compared to those without CHIP. Significant results are highlighted as follows: ** p < 0.01, * p < 0.05, ^†^ p < 0.10.

| Outcome | CHIP+<br>(univariable) |  | CHIP+<br>(adjusted for age, sex, and baseline eGFR) |  |
| --- | --- | --- | --- | --- |
| | $\beta \pm SE$ | P | $\beta \pm SE$ | P |
| hemoglobin (g/dL) | **$-1.5 \pm 0.4^{**}$** | $< 0.001$ | **$-1.2 \pm 0.4^{**}$** | 0.002 |
| MCV (fL) | $1.8 \pm 1.2$ | 0.15 | $1.1 \pm 1.4$ | 0.37 |
| urea (mmol/L) | **$4.4 \pm 1.8^{*}$** | 0.02 | $2.1 \pm 1.4$ | 0.12 |
| bicarbonate (mEq/L) | **$-2.6 \pm 0.7^{**}$** | $< 0.001$ | **$-2.0 \pm 0.6^{**}$** | 0.003 |
| PTH<sup>a</sup> (pg/mL) | **$6.7 \pm 2.6^{**}$** | 0.01 | **$4.6 \pm 2.2^{*}$** | 0.04 |
| phosphate (mmol/L) | $0.121 \pm 0.07^{\dagger}$ | 0.07 | $0.07 \pm 0.06$ | 0.26 |
| calcium (mmol/L) | $-0.06 \pm 0.03^{\dagger}$ | 0.07 | $-0.05 \pm 0.03$ | 0.13 |
| ferritin (μg/L) | **$115.0 \pm 37.1^{**}$** | 0.003 | **$101.5 \pm 37.2^{**}$** | 0.008 |
| transferrin saturation (%) | $2.9 \pm 2.7$ | 0.29 | $3.5 \pm 2.7$ | 0.20 |
| log(CRP) | $-0.04 \pm 3.2$ | 0.91 | $-0.09 \pm 3.3$ | 0.80 |
| log(FGF23) | $-0.05 \pm 0.11$ | 0.63 | $-0.06 \pm 0.09$ | 0.50 |

<sup>a</sup> PTH values exclude two individuals who had primary hyperthyroidism (one individual with CHIP who was untreated at baseline and one without CHIP who had received parathyroidectomy prior to baseline assessment).

Baseline PTH was significantly higher in the group with CHIP (18.8 ± 2.6 *vs*. 12.2 ± 1.2), and its association with CHIP status remained when adjusting for kidney function, age and sex (Table 2). Moreover, those with CHIP were significantly more likely to be on calcitriol for secondary hyperparathyroidism (32% vs. 5%; **χ**^2^ (3, *N* = 85) = 11.4, *P* = 0.0007). While they were not associated with dichotomous CHIP status (Table 2), log(FGF23) and total calcium were correlated with VAF when adjusting for age, sex, and baseline eGFR (Supplemental Table 2). Finally, serum bicarbonate levels were significantly lower in the group with CHIP (23.9 ± 0.5 vs. 26.5 ± 0.4), and this association also persisted when adjusting for kidney function, age and sex.

## Discussion

In a single-centre longitudinal cohort of 87 individuals with advanced CKD, the presence of CHIP in circulating blood cells was associated with lower baseline kidney function, higher rates of kidney functional decline and greater prevalence of CKD complications. Those with CHIP continued to have significantly lower hemoglobin concentrations over the follow-up period despite significantly greater prevalence of ESA use. Ferritin levels were significantly higher in those with CHIP but adding ferritin as a covariate to the regression model only accounted for part of the association between CHIP and lower hemoglobin. Anemia in CKD is the result of a heterogeneous mix of etiologies including but not limited to erythropoietin deficiency, chronic inflammation (IL-6-mediated hepcidin elevation), iron, B12 and folate deficiency.^24^ Inflammation observed in the CHIP state may contribute to the observed burden of anemia in CKD and may be targetable using new agents in development for anemia of inflammation in those with kidney disease.^25^ Since CHIP mutations affect hematopoietic stem cells directly, erythrocyte development and function may also be negatively affected^26^. It is conceivable that in CKD, CHIP adds a layer of stress to erythropoiesis that manifests as a quantitative defect with lower hemoglobin and increased mean red cell volume.^27^ While CHIP is not typically associated with anemia in surveys of healthy population cohorts,^28^ CHIP variants have been reported at a higher-than-expected prevalence in individuals with anemia.^29^

In summary, our study demonstrated an association between CHIP and CKD as well as an association between CHIP and the complications of CKD. In our cohort of patients with advanced CKD (eGFR < 60), 23% had CHIP, which is similar to other cohorts evaluating CHIP using targeted sequencing in the general population.^7,30,31^ More definitive estimates of the epidemiology of CHIP in CKD are expected to come from larger cohorts. Despite the limited sample size of 87 participants, given the comprehensive lab testing and serial follow up with 2,091 SCr measurements over 527 person-years, we were able to detect an association with incident decline in kidney function and common complications of CKD. Altogether, defining the relationship between CHIP and kidney health is important since CHIP is common with aging and strategies aimed at the surveillance, prevention and treatment of CHIP are in development.

## Data Availability

N/A

## Acknowledgements

We wish to thank Dr. Samuel Silver and Dr. Andrew Paterson for helpful suggestions for eGFR slope analyses and mixed model analyses, Dr. Alexander Bick, Dr. Pradeep Natarajan and, Seyedeh (Maryam) Zekavat for helpful discussions, as well as Dr. Xiao (Nick) Zhang for sample processing. This work was funded by a PSI Foundation Grant awarded to CV and MJR. MBL is supported by a Kidney Research Scientist Core Education and National Training (KRESCENT) Program New Investigator Award (cosponsored by the Kidney Foundation of Canada, the Canadian Society of Nephrology, and Canadian Institutes of Health Research).

## Declaration of conflicting interests

Unrelated to this work, MBL has received compensation as a speaker and advisory board member for Otsuka, Sanofi Genzyme, Reata, and Bayer.

## Notes

### Competing Interest Statement

The authors have declared no competing interest.

### Author Declarations

The study was approved by Queens University Health Sciences and Affiliated Teaching Hospitals Research Ethics Board (file 6023740).

