## Supplemental Tables for "Clonal hematopoiesis of indeterminate potential is associated with worse kidney function and anemia in a cohort of patients with advanced chronic kidney disease"

**Supplemental Information**

TABLE OF CONTENTS

1. Supplemental Table 1 – Characteristics of CHIP variants observed.

2. Supplemental Table 2 – CHIP associations with variant allele fraction (VAF).

**Supplemental Table 1 – Characteristics of CHIP variants observed**.

| Age* | Sex | Gene | Variant | VAF | Presumed etiology of CKD |
|---|---|---|---|---|---|
| 81-85 | M | *SF3B1* | p.Lys700Glu | 35.1 | unknown |
| | | *ASXL1* | p.Ile919Met | 21.3 | |
| 81-85 | F | *TET2* | p.Ala1769GlnfsTer22 | 3.1 | renovascular disease |
| 81-85 | M | *TET2* | p.Asp1376Tyr | 30.2 | renovascular disease & diabetes |
| | | *SETBP1* | p.Arg626Ter | 27.5 | |
| 81-85 | F | *DNMT3A* | p.Arg882His | 6.3 | renovascular disease |
| | | *DNMT3A* | p.Arg326Ser | 3.1 | |
| 81-85 | F | *TET2* | p.Tyr1295LeufsTer81 | 15.5 | renovascular disease & diabetes |
| | | *ASXL1* | p.Glu635ArgfsTer15 | 2.6 | |
| 81-85 | F | *TET2* | p.Cys1221Phe | 6.1 | renovascular disease |
| 76-80 | M | *DNMT3A* | p.Arg326Cys | 4.7 | obstructive nephropathy |
| 71-75 | F | *TET2* | p.Thr1975HisfsTer39 | 22.5 | obstructive nephropathy |
| 71-75 | F | *TET2* | p.Asp1384Gly | 5.2 | hypertension & diabetes |
| 71-75 | M | *ASXL1* | p.Arg1415Ter | 15.0 | interstitial nephritis |
| 71-75 | F | *TET2* | p.Arg1465Ter | 4.5 | IgA nephropathy |
| 66-70 | M | *STAG2* | p.Asp659Asn | 15.0 | renovascular disease |
| 60-65 | F | *DNMT3A* | p.Arg635Trp | 3.2 | membranous nephropathy |
| 60-65 | F | *DNMT3A* | p.Ile715ProfsTer65 | 4.1 | diabetes |
| 60-65 | M | *DNMT3A* | p.Arg729Trp | 5.9 | hypertension & diabetes |
| 60-65 | M | *TET2* | p.Val1213Gly | 10.9 | renovascular disease |
| 60-65 | M | *DNMT3A* | p.His694Tyr | 4.8 | renal artery stenosis |
| 46-50 | M | *DNMT3A* | p.Glu733Ter | 4.7 | renovascular disease & diabetes |
| 41-45 | M | *DNMT3A* | p.Gly421AlafsTer230 | 5.2 | polycystic kidney disease |
| | | *NF1* | p.Leu1187Arg | 3.6 | |
| 36-40 | M | *TET2* | p.Ala1505Thr | 43.0 | renovascular disease |

*Age range provided for de-identification purposes.

**Supplemental Table 2 – CHIP associations with VAF.** The results of univariable (VAF) and multivariable (VAF, age, sex, baseline eGFR) linear regressions are presented for each outcome of interest. The β coefficient indicates the quantitative change in the outcome per 1% increase in VAF. For those without CHIP, VAF was assigned a value of 0%. Significant results are highlighted as follows: ** $p < 0.01$, * $p < 0.05$, † $p < 0.10$.

| **Outcome** | **VAF** (univariable) | | **VAF** (adjusted for age, sex and baseline eGFR) | |
|---|---|---|---|---|
| | β | P | β | P |
| hemoglobin (g/dL) | **-0.07 ± 0.03*** | 0.02 | -0.05 ± 0.03† | 0.06 |
| MCV (fL) | **0.13 ± 0.07*** | 0.05 | 0.11 ± 0.07† | 0.10 |
| urea (mmol/L) | 0.14 ± 0.10 | 0.19 | 0.01 ± 0.08 | 0.89 |
| bicarbonate (mEq/L) | **-0.12 ± 0.04**** | 0.004 | **-0.09 ± 0.04*** | 0.02 |
| PTH[a] (pg/mL) | **0.29 ± 0.15*** | 0.05 | 0.15 ± 0.12 | 0.21 |
| phosphate (mmol/L) | -0.001 ± 0.004 | 0.86 | -0.003 ± 0.003 | 0.37 |
| calcium (mmol/L) | **-0.005 ± 0.002**** | 0.008 | **-0.004 ± 0.002*** | 0.02 |
| ferritin (µg/L) | **4.5 ± 2.1*** | 0.04 | 3.3 ± 2.1 | 0.12 |
| transferrin saturation (%) | 0.18 ± 0.15 | 0.24 | 0.17 ± 0.15 | 0.26 |
| log(CRP) | -0.02 ± 0.02 | 0.32 | -0.02 ± 0.02 | 0.33 |
| log(FGF23) | -0.003 ± 0.006 | 0.58 | **-0.009 ± 0.005*** | 0.05 |
